# The Potential Public Health Benefit of mRNA-1010 for Influenza Among Older Adults ≥65 Years in the United States

**DOI:** 10.64898/2026.09.20.26363308

**Authors:** Kelly Fust, Ekkehard Beck, Michele Kohli, Shannon Cartier, Milton Weinstein, Nicolas Van de Velde, Keya Joshi

## Abstract

**Background/Objectives:** Older adults are at high risk of influenza-related severe disease and complications. HD-IIV, currently recommended for adults ≥65 years, is egg-based and may be subject to egg-adapted mutations. An mRNA-based vaccine for influenza, mRNA-1010, has been approved by the Food and Drug Administration. Given the clinical benefits observed in mRNA-1010 clinical trials and potential to avoid egg-adapted mutations, this analysis explores the clinical impact of mRNA-1010 compared to HD-IIV in those ≥65 years in the United States.

**Methods:** This study used a decision-analytic model of influenza infection and downstream outcomes to project influenza burden with each vaccine over a one-year timeframe. Vaccine coverage, HD-IIV vaccine effectiveness (VE) and influenza epidemiology was estimated based upon Centers for Disease Control and Prevention data. The relative VE (rVE) for mRNA-1010 vs. HD-IIV was modelled based on data from the clinical trial and published literature on the impact of egg adaptation.

**Results:** The base-case analysis suggests mRNA-1010 may prevent an additional 232,200 symptomatic influenza infections, 130,000 outpatient cases, 21,100 influenza-related hospitalizations, and 2,200 influenza-related deaths compared to HD-IIV. The number needed to vaccinate to prevent one hospitalization was 404 for mRNA-1010 vs no vaccine and 514 for HD-IIV versus no vaccine, for an incremental NNV of 110. Scenario and deterministic sensitivity analyses results indicate that clinical outcomes prevented by mRNA-1010 are most sensitive to influenza incidence and VE.

**Conclusions:** mRNA-1010 may improve protection against influenza in US adults aged ≥65 years, leading to meaningful reductions in the clinical influenza-related disease burden in older adults.

## Introduction

Older adults ≥65 years continue to be at highest risk of severe disease following influenza infection. Data from the United States (US) Centers for Disease Control and Prevention (CDC) indicate that 50-70% of seasonal influenza-related hospitalizations and 70-85% of influenza-related deaths occur in adults aged ≥65 years.(1) This increased risk of severe illness in older adults is largely driven by the presence of chronic comorbidities and immunosenescence (i.e., age-related decline immune system functioning). Adults ≥65 years are also at increased risk of influenza-driven complications, with studies showing an increased risk of cardiovascular events and functional declines following an influenza infection.(2–4) The Advisory Committee on Immunization Practices (ACIP(5)) has recommended that vaccines that illicit a stronger immune response be preferentially used in adults ≥65 years in the US; therefore, for the 2025-2026 season, the CDC preferentially recommended trivalent versions of Fluzone® High-Dose (Sanofi Pasteur, Inc.) (HD-IIV), Fluad® (Seqirus Inc.) (aIIV4), or Flublok® (Protein Sciences Corporation)(RIV), for adults ≥65 years.(1, 6)

An mRNA-based vaccine for influenza, mRNA-1010 (mFLUSIVA®, Moderna, Inc.), is licensed by the FDA with an indication in adults aged ≥50 years; mRNA-1010 was approved by the US Food and Drug Administration (FDA) in August of 2026 and is available for the 2026-2027 influenza season.(7–10) The efficacy, immunogenicity, and safety of mRNA-1010 has been evaluated in Phase 3 studies mRNA-1010-P303 (NCT05827978) and mRNA-1010-P304 (NCT06602024). mRNA-1010-P304, is a Phase 3, randomized, double-blind, active-controlled clinical trial comparing mRNA-1010 to IIV3 (Fluzone®; Sanofi Pasteur, Inc.) in medically stable adults aged ≥50 years.(11) The clinical trial primary endpoint was the relative vaccine efficacy (rVE) of mRNA-1010 versus a standard dose comparator to prevent the first episode of reverse transcription polymerase chain reaction (RT-PCR)-confirmed protocol-defined influenza-like illness (ILI) caused by any influenza A or B strains over the time period beginning 14 days after study vaccination through the end of the influenza season.(12) The rVE of mRNA-1010 compared to standard dose influenza vaccine was 26.6% (95% CI: 16.7%, 35.4%) among adults ≥50 years. In the subgroup of adults aged ≥65 years, the rVE for mRNA-1010 was 27.4% (95% CI: 12.1%, 40.0%) compared to standard dose influenza vaccine.(12) An additional three-part trial, mRNA-1010-P303 (P303), evaluated the immunogenicity, reactogenicity, and safety of quadrivalent mRNA-1010 in adults aged ≥18 years vs licensed standard (for adults 18-64 years)- and high-dose (for adults ≥65 years) quadrivalent seasonal influenza vaccines. Results from P303 Part C found mRNA-1010 met statistical superiority criteria compared to Fluzone®-HD across all 8 co-primary immunogenicity endpoints defined by geometric mean titers (GMT) and seroconversion rates (SCR).(11, 12) The American Academy of Family Physicians (AAFP) includes mRNA-1010 as a preferentially recommended enhanced product (in addition to high-dose, adjuvanted, and combination vaccines) for adults aged ≥65 years for the 2026-2027 influenza season.(10)

Two of the vaccines currently recommended by ACIP and the AAFP for preferential use in adults ≥65 years, HD-IIV and aIIV, are egg-based vaccines, meaning they are produced by allowing the virus to replicate within chicken eggs.(10, 13) During the incubation of the virus, it may develop a mutation that allows the virus to replicate itself more efficiently. If these egg-adaption mutations affects the antigenic match to circulating viruses, VE may be reduced. In contrast, mRNA-1010 is manufactured using *in vitro* techniques and therefore is not subject to egg adaptation. In the United Kingdom (UK), the Joint Committee on Vaccination and Immunisation (JCVI) preferentially recommends the use of influenza vaccines that are not egg-based for those ages 18 to 64 years given concerns about the impact of egg-adaptation on effectiveness.(14)

Given the clinical benefits observed in mRNA-1010 clinical trials, the potential to avoid egg adapted mutations with mRNA-1010, and the severe influenza-related disease burden in older adults, the aim of this analysis is to project the clinical impact of mRNA-1010 compared to the egg-based HD-IIV influenza vaccine, which are both recommended as preferred by the AAFP for adults ≥65 years of age in the United States.

## Methods

### Overview

In this analysis, vaccination with mRNA-1010 is compared to vaccination with HD-IIV among US adults ≥65 years. Although aIIV4 is also an egg-based vaccine, HD-IIV was selected as the model comparator as it is a longstanding market leader in vaccinations for older adults in the US. Further, following a systematic review, ACIP indicated that while there are VE studies directly comparing HD-IIV3 and aIIV, they concluded that there are no significant differences in VE across seasons.(5, 15)

This study used a decision-analytic model of influenza infection and its downstream outcomes, capturing only the direct benefits of vaccination.(16) This conservative static approach is consistent with the CDC in their annual estimates of the burden of influenza in the US.(17–20)

Analyses were performed over a one-year timeframe. The model was used to project the following clinical outcomes: numbers of symptomatic infections, outpatient treated cases, hospitalizations, and deaths. The numbers needed to vaccinate (NNV) in order to prevent one symptomatic infection, one hospitalization, and one death were also calculated for each vaccine to estimate the incremental NNV of mRNA-1010 vs HD-IIV (e.g., the number of additional persons that would need to be vaccinated to prevent one influenza outcome with HD-IIV vs mRNA-1010).

### Model Structure

A Markov model with a Well and a Dead health state was used to track the target population on a monthly basis over a one-year time horizon. The cohort begins in the Well state, and each month a portion may receive an influenza vaccine according to observed monthly vaccine coverage rates. In addition, each month, individuals in the Well health state face the risk of symptomatic influenza infection, which is dependent on infection incidence and their vaccination status. Infection incidence varies by age group, while vaccines reduce the probability of infection and may incrementally reduce the probability of hospitalization following infection. Each vaccine was assigned an initial VE but, because the effectiveness of the influenza vaccines wanes over time, VE was assumed to decline linearly on a monthly basis.(21) As VE for both the egg-based vaccine and the mRNA vaccine wane over time but the proportion vaccinated increases during the course of the influenza season, the average monthly VE is a function of the fraction of the age group vaccinated each month and the vaccine effectiveness. (See Supplemental material). If persons develop an infection, they move through the influenza infection tree (Figure 1), and the health outcomes and associated costs are counted. At the end of the influenza infection tree in each monthly cycle, individuals either return to the Well health state or move into the Dead health state to start the next Markov cycle. Those who do not develop an infection remain in the Well health state.

**Figure 1.**
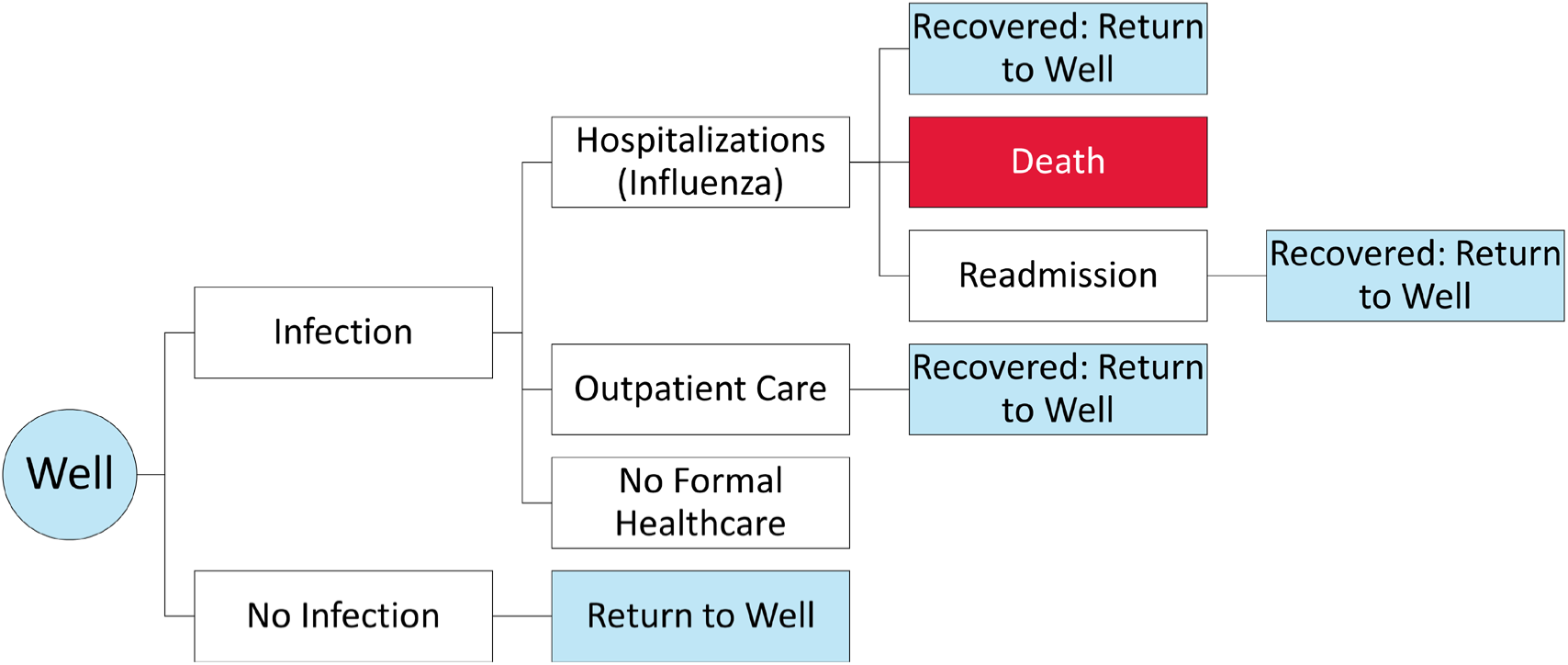
Structure of the influenza infection tree.

### Model Inputs: Population Size

Results are reported for the target population of all adults aged ≥65 years. The size of this population was estimated to be 61,179,918 based on the CDC burden of illness model data (2024–2025).(17)

### Model Inputs: Incidence of infection

For the base-case analysis, we chose to focus on 9 influenza seasons from 2011-2012 to 2019-2020 because Russel et al(7) previously reviewed US data to determine which of these seasons were impacted by egg adaptation. The model therefore required the monthly incidence of symptomatic infection without vaccination for 2011/12 to 2019/20. An annual incidence of 7.3% was calculated by summing the average number of symptomatic infections(17) and the average prevented infections(18)outputs from the CDC burden of illness model for the years 2011/12 to 2019/20 and then dividing by the average population size to calculate a rate (see Supplemental material). As the model requires monthly estimates of incidence of symptomatic infection, but monthly data are available only for hospitalized infections, the monthly rates of hospitalization from the influenza hospitalization surveillance network (FluSurv-NET) for 2011/12 to 2019/20(22) were averaged together and the monthly pattern of this average was used to distribute the annual incidence of infection without vaccination by month. To ensure that the model reproduced numbers similar to the CDC burden of illness, a validation was done using the average target population size from the seasons of interest (see Supplemental material).

As the incidence of influenza varies widely from year to year, a number of sensitivity analyses were conducted. A scenario analysis was conducted using data from two post-COVID-19 pandemic influenza seasons: 2023-2024 and 2024-2025, reflecting the seasons the P303 and P304 studies were conducted. In addition, the data from the single season of 2017-2018, during which vaccine VE was impacted by egg adaptation and incidence of infection was high, was used to generate a “worst-case scenario” for seasonal influenza. Deterministic sensitivity analyses (DSAs) were also conducted by varying incidence using the high and low season incidence from the 9 seasons averaged for the base-case analysis. Therefore, the data from 2014/15 were used to create a high incidence scenario (annual incidence:12.4%), while the data from 2011/12 were used to create a low incidence scenario (annual incidence: 3.2%). The incidence scenarios therefore varied in terms of both magnitude and the month when they peaked. The base case peak was in January, the high scenario peak was in December, and the low scenario peak was in March. (See Supplemental material for the monthly incidence.)

### Model Inputs: Vaccine coverage

The portion unvaccinated is the same in both strategies and is estimated as an average of the observed vaccine coverage rates (VCRs) for the 2011/12 to 2019/20 seasons as reported by the CDC(23, 24) (See Table in the Supplemental material). The average cumulative vaccine coverage for those ≥65 years and older was 65.4%. In deterministic sensitivity analyses, the absolute incidence of vaccine coverage was varied by 5%.

### Vaccine effectiveness

Similar to DeLuca et al., VE across a number of pre-pandemic seasons was used in this analysis.(25) The seasonal VE of influenza vaccines in adults ≥65 years was estimated in the CDC burden of illness model(18) for the nine seasons reviewed by Russell et al.(7) (2011/12 – 2019/20), and ranged from 12% to 50%; these were used to estimate initial VE values as described in the Supplemental material. The average of these initial VE values was assumed to represent the base case VE for HD-IIV, while the range was used in deterministic sensitivity analyses.

Precise antigenic matching to selected influenza vaccine strains and the non-egg based manufacturing process both have the potential to improve VE compared to currently used influenza vaccines.(7, 8) Therefore, the incremental VE for mRNA-1010 comprises two components: (1) modeled relative VE (rVE) of mRNA-1010 vs. HD-IIV, reflecting the improvement in immunogenicity, and (2) rVE due to the ability of an mRNA vaccine to avoid egg-adapted mutations.

The first component was estimated using a previously established correlate of protection (CoP) modeling approach(26) to generate the predicted rVE for mRNA-1010 vs HD-IIV using immunogenicity data (measured as hemagglutination inhibition (HAI) geometric mean titres (GMTs)) from P303 Part C. A sigmoidal function, based on the model described by Dunning et al. (2016)(26) was first used to model the relationship between the strain-specific HAI GMTs and vaccine efficacy using data from the P304 trial.(27) The model was then used on the immunogenicity data from the -P303 Part C trial to project a seasonal rVE for mRNA-1010 compared to HD-IIV of 12.4% (95% CI: 3.4% - 21.3%).(27)

The second component of rVE reflected the non-reliance on egg-based manufacturing. During the years the Phase 3 trials for mRNA-1010 were conducted, CDC and World Health Organization (WHO) analyses generally found that vaccine antigens for A(H1N1)pdm09, A(H3N2), and B/Victoria elicited antibodies that reacted well with circulating influenza viruses, or that reduced antigenic match was attributable primarily to antigenic drift rather than egg adaptation.(28–31) However, during the 2023– 2024 season, there was some variable evidence suggesting that egg-adaptive mutations in the A(H1N1)pdm09 vaccine virus may have contributed to reduced antigenic match or vaccine effectiveness against certain circulating H1N1 viruses, although the clinical significance of these mutations and their contribution relative to other factors remain uncertain.(28, 29, 32, 33) Therefore, in years where severe egg adaptation occurs, the rVE of mRNA-1010 vs. HD-IIV is expected to be higher than observed in P303.

Russell et al(7) found evidence that egg adaptation reduced VE in Northern hemisphere vaccines in 6 of the 9 influenza seasons (66.7%) between 2011-2012 to 2019-2020. Stein et al. conducted a retrospective test-negative design to compare the use of Flucelvax® (Seqirus Inc.) (ccIIV), which is replicated in mammalian cells, to standard egg-based vaccines (IIV) across three influenza seasons in the US.(32) They found that the rVEs of ccIIV versus IIV in the 2017-18, 2018-19, and 2019-20 seasons were 14.8% (95% confidence interval [CI]: 7.0%-22.0%), 12.5% (95% CI: 4.7%-19.6%), and 10.0% (95%CI: 2.7%-16.7%), respectively. The rVE for an egg-based vaccine compared to a non-egg-based vaccine (rVEea) in one season was estimated as 12.4% based on the 3-season average from this study by Stein et al.(32) The rVEea was assumed to apply to only 66.7% of seasons based on Russell et al.(7) and was therefore reduced to 8.3% (12.4% X 66.7%). The impact of egg adapted mutations on seasonal was varied from 12.4% in deterministic sensitivity analyses (DSAs) from 2.7% to 22.0% based on the range of the 95% CIs for the 3 seasons in the Stein study.(32) The frequency was also varied from 66.7% in DSAs where we assumed egg adaptation occurs in 50% to 75% of seasons.

The rVE from the CoP model and the rVE for egg-adaptation were considered to be independent and were combined together, as follows, for the base case. As Stein et al.(32) conducted their analysis of the impact of egg adaptation with logistic regression, their rVE was calculated as (1 – odds ratio) X 100%. In order to combine this benefit with the rVE calculated by the correlation of protection model, it was assumed that the odds ratio approximates a relative risk. The overall rVE of mRNA-1010 versus HD-IIV was therefore 19.6% in base case. Scenario analyses were conducted with rVE based on CoP data only and rVE based on egg-adaptation only. An additional scenario analysis was performed assuming the rVE from the CoP model was replaced with 6.5% (calculated from Izurieta et al.(34)) estimated based on the difference in rVE between RIV and IIV versus HD and IIV.

Once calculated, the initial VE is applied in the influenza infection tree to the portion of the cohort newly vaccinated each month. Given the rVEs presented above, for the analyses conducted with data from 2011/12 to 2019/20, the absolute increase in VE for mRNA-1010 compared to HD-IIV was 5.3% (CoP results alone), 3.6% (lack of egg-adaptation alone), and 8.5% (CoP results and lack of egg-adaptation). As a conservative assumption, we assume that none of the vaccines have an independent effect on hospitalizations;(15) therefore the VE against hospitalization was set equal to the VE against infection (i.e. no incremental effect). The monthly waning rate for both the high-dose and standard dose vaccines was assumed to be 8% per month.(21, 35)

### Additional probabilities from the influenza decision tree

Those who develop symptomatic influenza infection may not seek formal care, may be treated as an outpatient only (including either outpatient physician visits or emergency department visits without hospital admission), or may be admitted to the hospital. The age-specific probabilities of hospitalization, outpatient visits, and no formal care were estimated consistently with DeLuca et al. (2023)(25) based on US CDC Burden of Illness data as 9.1%, 56.0% and 34.9% respectively.

Age-specific estimates of in-hospital mortality were also estimated from the CDC burden of illness model(17) for 2011/2012 to 2019/2020 as 10.6%. Given that the CDC uses death certificate data and statistical modelling to capture mortality from influenza, which includes influenza-related deaths that occur following discharge, separate estimates of post-discharge mortality are not included in the model. Finally, influenza-related mortality is assumed to apply to hospitalized patients only; patients surviving the initial hospitalization are subject to the risk of readmission (8.76%). Estimates are provided in Table 1.

**Table 1.** Model Parameters.

| Parameter | Value (Range) |  |  | Source |
| --- | --- | --- | --- | --- |
| Decision Tree Probabilities |  |  |  |  |
|  | % Hospitalized | % Outpatient Care | % No Formal Care |  |
| Distribution of care following symptomatic infection | 9.1% (±25%) | 56.0% | 34.9% (±25%) | US CDC Preliminary Estimated Flu Disease Burden(17, 18) |
| In-hospital mortality | 10.6% (9.1%, 17.9%) |  |  | Calculated using CDC BOI data (2011/2012 - 2019/2020)(17, 18) |
| Percentage readmitted (of those hospitalized) | 8.76% (±25%) |  |  | Optum Claims Database Analysis |

BOI: Burden of illness; CDC: Centers for Disease Control and Prevention; US: United States

### Analysis of uncertainty: Sensitivity Analyses

Multiple sensitivity analyses were performed to assess the impact of key model inputs on clinical outcomes. Several scenario analyses, described in the text above, were conducted including ones for incidence and vaccine effectiveness. Deterministic sensitivity analyses (DSAs) were conducted to understand the important drivers of the results and were summarized in tornado diagrams. The DSAs for the parameters related to influenza incidence, vaccine coverage, initial VE, probability of egg adaptation occurring, and the rVE of mRNA-1010 versus HD-IIV (ranges from the COP model and egg adaptation) were summarized in the sections above. Infection consequence probabilities (i.e., hospitalization and mortality) were also included in DSAs. Estimates were varied according to 95% confidence intervals (CIs) or reported ranges where available; for all other DSAs, parameters were varied by ±25% of their base-case value (Table 1).

## Results

As presented in Table 2, vaccination with mRNA-1010 was estimated to avert an additional 232,000 symptomatic influenza infections, 130,000 outpatient cases, 21,100 influenza-related hospitalizations, and 2,200 influenza-related deaths compared to vaccination with HD-IIV. The corresponding number of additional doses with HD-IIV required compared to mRNA-1010 to avert one symptomatic influenza infection, outpatient episode, hospitalization or death for mRNA-1010 compared to HD-IIV were estimated at 10, 18, 110, and 1,035 respectively (Table 3). In order for HD-IIV to yield the same number of symptomatic infections, hospitalizations, and deaths as mRNA-1010, the overall vaccine coverage rate would have to increase to 83.2% in those ages ≥65 years (representing an increase of 17.8% compared to the base-case annual coverage of 65.4%).

**Table 2.** Base-Case Clinical Outcomes Results.

| Strategy | Health outcomes |  |  |  |
| --- | --- | --- | --- | --- |
|  | Symptomatic Infection | Outpatient Treatment Only | Hospitalization | Deaths |
| HD-IIIV | 3,604,272 | 2,018,392 | 327,661 | 34,661 |
| mRNA-1010 | 3,372,116 | 1,888,385 | 306,556 | 32,429 |
| Outcomes averted by mRNA-1010 | 232,155 | 130,007 | 21,105 | 2,233 |

**Table 3.**
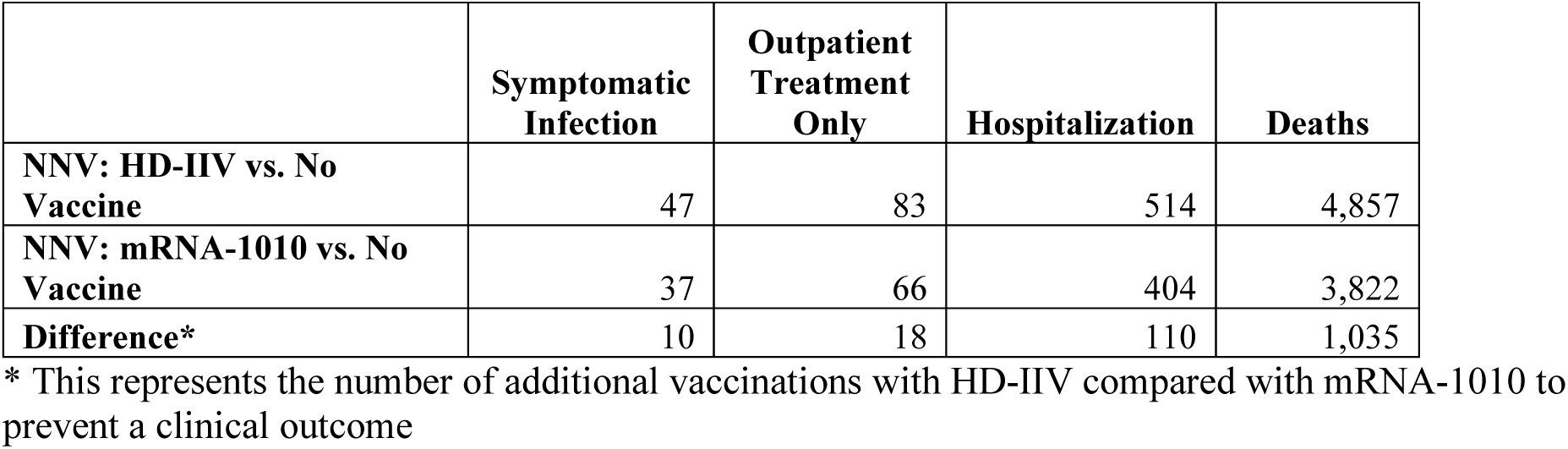
Number Needed to Vaccinate (NNV)

|  | Symptomatic Infection | Outpatient Treatment Only | Hospitalization | Deaths |
| --- | --- | --- | --- | --- |
| **NNV: HD-IIIV vs. No Vaccine** | 47 | 83 | 514 | 4,857 |
| **NNV: mRNA-1010 vs. No Vaccine** | 37 | 66 | 404 | 3,822 |
| **Difference\*** | 10 | 18 | 110 | 1,035 |

\* This represents the number of additional vaccinations with HD-IIIV compared with mRNA-1010 to prevent a clinical outcome

In scenario analyses, changing the rVE had the most impact. For example, utilizing the rVE from the COP model only yields 146,562 symptomatic infections, 13,324 hospitalizations, and 1,409 deaths averted by mRNA-1010 (Technical Appendix Table 5). Utilizing only the rVE associated with egg adaptation from the base case (i.e., 8.27%) yields 97,707 symptomatic infections, 8,882 hospitalizations, and 940 deaths averted by mRNA-1010 (Technical Appendix Table 5. In a scenario analysis using only the rVE for egg adaptation assuming a probability that egg adaptation occurs in one third of seasons (half the probability of base case), mRNA-1010 averted 48,853 symptomatic infections, 4,441 hospitalizations, and 470 deaths; (Technical Appendix Table 5. Utilizing an rVE of 6.5% yields 168,184 symptomatic infections, 15,289 hospitalizations, and 1,617 deaths averted by mRNA-1010. Results of scenario analyses examining incidence across various influenza seasons are also presented in the Technical Appendix (Table 5. Overall, varying the years included to calculate the average incidence without influenza vaccination had a small impact on the results overall. For 2017-2018, which represents a season with high incidence where the probability of egg adaptation occurring is 100%, the clinical cases prevented by mRNA-1010 are predicted to double.

Results of deterministic sensitivity analyses (Figure 3) indicate that clinical outcomes prevented by mRNA-1010 are most sensitive to influenza incidence, rVE from CoP model, and the VE of HD-IIV. Incidence has the greatest impact on clinical outcomes averted by mRNA-1010 compared to HD-IIV. Influenza incidence reflective of the 2014/2015 season (“high incidence”) increases symptomatic infections, hospitalizations, and death prevented by mRNA-1010 by 67% (compared to the base-case) to 387,639, 35,240, and 3,728, respectively. Varying influenza incidence to the 2011/2012 season (“low incidence”) decreases symptomatic infections, hospitalizations, and deaths prevented by 55% to 105,722, 9,611, and 1,017, respectively. Varying the rVE based on the COP model to the upper bounds of the 95% CI yields 328,657 symptomatic infections prevented by mRNA-1010 and 29,878 and 3,161 hospitalizations and deaths prevented, respectively (38% increase), while varying the rVE to the lower bounds of the 95% CI yields 134,572 symptomatic infections prevented by mRNA-1010 and 12,234 and 1,294 hospitalizations and deaths prevented, respectively (43% decrease). Varying the VE of HD-IIV to the upper and lower estimates over the last 9 seasons yields 140,240 and 304,367 symptomatic infections, 12,749 and 27,670 hospitalizations, and 1,349 and 2,297 deaths prevented by mRNA-1010, respectively. The number of deaths prevented is also sensitive to the estimate of mortality, while the number of hospitalizations and death prevented are also sensitive to the probability of hospitalization. Detailed results are presented in the Technical Appendix (Table 6.

**Figure 3.**
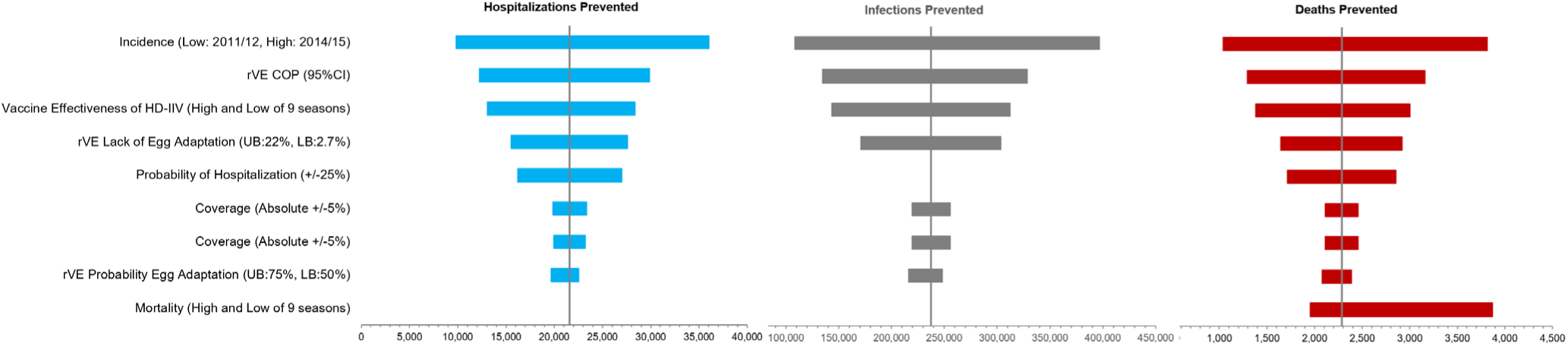
Deterministic Sensitivity Analyses (Clinical Results) COP: correlate of protection; rVE: relative vaccine effectiveness

## Discussion

Results of the base-case analysis, including both the clinical benefit of mRNA-1010 based on the COP model as well as lack of egg-adapted mutations, suggest compared to HD-IIV, mRNA-1010 may prevent an additional 232,200 symptomatic influenza infections, 130,000 outpatient cases, 21,100 influenza-related hospitalizations, and 2,200 influenza-related deaths. Results of scenario and deterministic sensitivity analyses indicate that clinical outcomes prevented by mRNA-1010 are most sensitive to influenza incidence and VE.

This analysis is subject to several limitations related to VE. The seasonal VE of influenza vaccines in adults ≥65 years was estimated based on the CDC burden of illness model(18) for 2011/12 – 2019/20, to estimate initial VE values which were assumed to represent the VE for HD-IIV. Although it is possible that VE for standard-dose vaccines may be represented in these estimates, by the 2019-2020 influenza season almost 90% of those aged ≥65 years were receiving enhanced vaccines.(34) Further, the average estimate considered in the analysis was in line with HD-IIV specific VE estimates from recent seasons (2023–2025) from the California Department of Public Health. (36–38)

VE assumptions used in base-case analyses for mRNA-1010 are likely conservative. The waning pattern of mRNA-1010 was assumed to be equivalent to HD-IIV; however, clinical trial data suggest that mRNA-1010 efficacy persisted throughout the full influenza season, and that HAI titers for mRNA-1010 over HD-IIV persisted over a period of 6 months.(11, 12) Additionally, rVE estimates from the COP model alone may be conservative as they do not capture the potential impact in a severe egg adapted season. Base case estimates of VE independently combining both the modeled rVE estimates and benefits associated with egg adaptation will need to be confirmed once real world data for mRNA-1010 become available. Further, the full benefits of mRNA-1010 may be underestimated due to the use of a static versus dynamic model structure (i.e., benefits associated with herd immunity are not reflected in the present analysis).[14]

The potential future clinical benefits of mRNA-1010, associated with the mRNA platform, have not been fully examined within this analysis. Each year, the Food and Drug Administration (FDA) mandates the viruses to be included in the seasonal influenza vaccines in the US(39) in February or March to allow manufacturers time to produce sufficient quantities of vaccines for the influenza vaccination campaigns, which begin in September. During the 6 month lag between March and September, the circulating viruses may become antigenically different from the FDA-selected vaccine viruses, or “mismatched”, and genetic drift is said to have occurred. In the 2025/26 season, for example, a new subclade of A/H3N2 (subclade J.2.4.1, also called subclade K), that is considered to be antigenically drifted from the A/H3N2 virus in the vaccine, emerged.(40) As mRNA-1010 can be manufactured more quickly than other influenza vaccines, the choice of virus for these vaccines may be delayed by 3 months until June, making it possible to reduce the probability of mismatch between the vaccine and circulating strains. In addition to using a genetic sequence approach that had a benefit in 51/63 seasons, delaying strain selection by 3 months could add benefit in an additional 14 seasons (41), which would improve both the clinical and economic benefits associated with mRNA-1010. This additional benefit, which is associated with the mRNA-platform, should be explored in future research.

Influenza incidence is variable across seasons; however, the average of incidence of the CDC burden of illness data aligns with the incidence reported by other sources.(42, 43) Additionally, while the CDC collects information on deaths from death certificates for hospitalizations reported in FluServ Net in order to capture mortality post-discharge, the follow-up period may be limited, and deaths post-discharge may disproportionally affect those at older ages.(44) While the estimated number of US influenza-associated deaths is reported annually, detailed data on the epidemiology of influenza-associated deaths, including the burden of in-hospital compared to post-hospital mortality are limited and may be underestimated(44); additionally, recent studies have found that mortality rates can remain elevated for longer periods following discharge.(45)

In conclusion, mRNA-1010 may improve protection against influenza in US adults aged ≥65 years, leading to reductions in the clinical influenza-related disease burden in this population compared to existing enhanced vaccines. The mRNA platform has the potential to further improve influenza vaccine effectiveness by reducing the time between strain choice and vaccine launch and this potential needs to be explored in future research.

## Author Contributions

Conceptualization, KF, MK and KJ; Methodology, KF, MK, KJ, MW, SC; Validation, MK and SC; Formal Analysis, KF, MK and SC; Writing – Original Draft Preparation, KF and MK; Writing – Review & Editing, KJ, EB, MW, NV, and SC; Funding Acquisition, KJ

## Funding

This study was funded by Moderna, Inc.

## Institutional Review Board Statement

Not applicable

## Informed Consent Statement

Not applicable

## Data Availability Statement

No new data were created or analyzed in this study. Data sharing is not applicable to this article.

## Acknowledgements

None

## Conflicts of Interest

MK is a shareholder in Quadrant Health Economics Inc, which was contracted by Moderna, Inc. to conduct this study. KF, MW and SC are consultants to Quadrant Health Economics Inc. KJ, EB, and NV are employed by Moderna, Inc. and may hold stock/stock options in the company.

## Notes

### Competing Interest Statement

This study was funded by Moderna, Inc.MK is a shareholder in Quadrant Health Economics Inc, which was contracted by Moderna, Inc. to conduct this study. KF, MW and SC are consultants to Quadrant Health Economics Inc. KJ, EB, and NV are employed by Moderna, Inc. and may hold stock/stock options in the company.

### Summary of Updates

The numbers needed to vaccinate in the abstract results section have been updated to reflect 404 for mRNA-1010 and 514 for HD-IIV.

## References

1. Center for Disease Control and Prevention. Flu and People 65 Years and Older [Available from: https://www.cdc.gov/flu/highrisk/65over.htm.

2. Kwong JC, Schwartz KL, Campitelli MA. Acute Myocardial Infarction after Laboratory-Confirmed Influenza Infection. N Engl J Med. 2018;378(26):2540–1.

3. Chow EJ, Rolfes MA, O’Halloran A, Anderson EJ, Bennett NM, Billing L, et al. Acute Cardiovascular Events Associated With Influenza in Hospitalized Adults : A Cross-sectional Study. Ann Intern Med. 2020;173(8):605–13.

4. Andrew MK, MacDonald S, Godin J, McElhaney JE, LeBlanc J, Hatchette TF, et al. Persistent Functional Decline Following Hospitalization with Influenza or Acute Respiratory Illness. J Am Geriatr Soc. 2021;69(3):696–703.

5. Grohskopf LA, Blanton LH, Ferdinands JM, Chung JR, Broder KR, Talbot HK, et al. Prevention and Control of Seasonal Influenza with Vaccines: Recommendations of the Advisory Committee on Immunization Practices - United States, 2022-23 Influenza Season. MMWR Recomm Rep. 2022;71(1):1–28.

6. Grohskopf LA, Blanton LH, Ferdinands JM, Reed C, Dugan VG, Daskalakis DC. Prevention and Control of Seasonal Influenza with Vaccines: Recommendations of the Advisory Committee on Immunization Practices - United States, 2025-26 Influenza Season. MMWR Morb Mortal Wkly Rep. 2025;74(32):500–7.

7. Russell CA, Fouchier RAM, Ghaswalla P, Park Y, Vicic N, Ananworanich J, et al. Seasonal influenza vaccine performance and the potential benefits of mRNA vaccines. Hum Vaccin Immunother. 2024;20(1):2336357.

8. Harding AT, Heaton NS. Efforts to Improve the Seasonal Influenza Vaccine. Vaccines (Basel). 2018;6(2).

9. United States Food and Drug Administration. Clinical Review - MFLUSIVA 2026 [Available from: https://www.fda.gov/media/194140/download.

10. American Academy of Family Physicians. 2026-2027 Influenza, RSV, and SARS-CoV-2 Immunization Recommendations 2026 [Available from: https://www.aafp.org/assets/image/upload/v1788277985/pdf_2026_through_2027_influenza_rsv_sars_covid_2_guidance.pdf.

11. Leroux-Roels I, Huang G, Ferguson M, Kohli A, Clark R, Bickel M, et al. Efficacy and Safety of an mRNA Seasonal Influenza Vaccine in Adults. N Engl J Med. 2026;394(18):1803–13.

12. Soens M, Ananworanich J, Hicks B, Lucas KJ, Cardona J, Sher L, et al. A phase 3 randomized safety and immunogenicity trial of mRNA-1010 seasonal influenza vaccine in adults. Vaccine. 2025;50:126847.

13. Center for Disease Control and Prevention. How Influenza (Flu) Vaccines Are Made [Available from: https://www.cdc.gov/flu/vaccine-process/.

14. Government of UK DoHaSC. JCVI statement on influenza vaccines for 2026 to 2027 2025 [Available from: https://www.gov.uk/government/publications/flu-vaccines-2026-to-2027-jcvi-advice-16-july-2025/jcvi-statement-on-influenza-vaccines-for-2026-to-2027.

15. Centers for Disease Control and Prevention. GRADE: Higher Dose and Adjuvanted Influenza Vaccines for Persons Aged 2024 [Available from: https://www.cdc.gov/acip/grade/influenza-older-adults.html.

16. Pitman R, Fisman D, Zaric GS, Postma M, Kretzschmar M, Edmunds J, et al. Dynamic transmission modeling: a report of the ISPOR-SMDM Modeling Good Research Practices Task Force--5. Value Health. 2012;15(6):828–34.

17. Centers for Disease Control and Prevention. Estimated US Flu Disease Burden [Available from: https://www.cdc.gov/flu-burden/php/data-vis/index.html.

18. Centers for Disease Control and Prevention. Flu burden prevented by vaccination [Available from: https://www.cdc.gov/flu-burden/php/data-vis-vac/index.html.

19. Reed C, Chaves SS, Daily Kirley P, Emerson R, Aragon D, Hancock EB, et al. Estimating influenza disease burden from population-based surveillance data in the United States. PLoS One. 2015;10(3):e0118369.

20. Rolfes MA, Foppa IM, Garg S, Flannery B, Brammer L, Singleton JA, et al. Annual estimates of the burden of seasonal influenza in the United States: A tool for strengthening influenza surveillance and preparedness. Influenza Other Respir Viruses. 2018;12(1):132–7.

21. Ferdinands JM, Gaglani M, Martin ET, Monto AS, Middleton D, Silveira F, et al. Waning Vaccine Effectiveness Against Influenza-Associated Hospitalizations Among Adults, 2015-2016 to 2018-2019, United States Hospitalized Adult Influenza Vaccine Effectiveness Network. Clin Infect Dis. 2021;73(4):726–9.

22. Centers for Disease Control and Prevention. Influenza hospitalization surveillance network (FLUSurv-NET) [Available from: https://www.cdc.gov/fluview/overview/influenza-hospitalization-surveillance.html#cdc_generic_section_1-flusurv-net.

23. Centers for Disease Control and Prevention. Influenza vaccination coverage for all ages (6+months) [Available from: https://data.cdc.gov/Flu-Vaccinations/Influenza-Vaccination-Coverage-for-All-Ages-6-Mont/vh55-3he6/about_data.

24. Centers for Disease Control and Prevention. Weekly influenza vaccination coverage and intent for vaccination, overall, by selected demographics and jurisdictions, among adults 18 years and older [Available from: https://data.cdc.gov/Flu-Vaccinations/Weekly-Influenza-Vaccination-Coverage-and-Intent-f/sw5n-wg2p/about_data.

25. DeLuca EK, Gebremariam A, Rose A, Biggerstaff M, Meltzer MI, Prosser LA. Cost-effectiveness of routine annual influenza vaccination by age and risk status. Vaccine. 2023;41(29):4239–48.

26. Dunning AJ, DiazGranados CA, Voloshen T, Hu B, Landolfi VA, Talbot HK. Correlates of Protection against Influenza in the Elderly: Results from an Influenza Vaccine Efficacy Trial. Clin Vaccine Immunol. 2016;23(3):228–35.

27. Nguyen VH SE, Youhanna J, et al. Estimating the Relative Vaccine Efficacy of mRNA-1010 to High-Dose Influenza Vaccine in Adults ≥65 Years Using a Correlate of Protection Framework. Vaccines. 2026;14(9):750.

28. Centers for Disease Control and Prevention. Influenza Activity in the United States during the 2023–2024 Season and Composition of the 2024–2025 Influenza Vaccine 2024 [Available from: https://www.cdc.gov/flu/whats-new/flu-summary-2023-2024.html.

29. World Health Organization. Recommended composition of influenza virus vaccines for use in the 2024-2025 northern hemisphere influenza season 2024 [Available from: https://cdn.who.int/media/docs/default-source/influenza/who-influenza-recommendations/vcm-northern-hemisphere-recommendation-2024-2025/recommended-composition-of-influenza-virus-vaccines-for-use-in-the-2024-2025-northern-hemisphere-influenza-season.pdf?sfvrsn=2e9d2194_7&download=true.

30. World Health Organization. Recommended composition of influenza virus vaccines for use in the 2025-2026 northern hemisphere influenza season 2025 [Available from: https://cdn.who.int/media/docs/default-source/influenza/who-influenza-recommendations/vcm-northern-hemisphere-recommendation-2025-2026/recommended-composition-of-influenza-virus-vaccines-for-use-in-the-2025-2026-northern-hemisphere-influenza-season.pdf?sfvrsn=857c2e9b_13&download=true.

31. Center for Disease Control and Prevention. Influenza Activity in the United States during the 2024–25 Season and Composition of the 2025–26 Influenza Vaccine [Available from: https://www.cdc.gov/flu/whats-new/2025-2026-influenza-activity.html.

32. Stein AN, Mills CW, McGovern I, McDermott KW, Dean A, Bogdanov AN, et al. Relative Vaccine Effectiveness of Cell-vs Egg-Based Quadrivalent Influenza Vaccine Against Test-Confirmed Influenza Over 3 Seasons Between 2017 and 2020 in the United States. Open Forum Infect Dis. 2024;11(5):ofae175.

33. Lucaccioni H, Pozo F, Perez-Gimeno G, Durrwald R, Uras M, Domegan L, et al. Vaccine effectiveness against medically attended, laboratory-confirmed influenza in the I-MOVE primary care network in Europe, VEBIS project, 2024/25. Expert Rev Vaccines. 2026;25(1):2645378.

34. Izurieta HS, Lu M, Kelman J, Lu Y, Lindaas A, Loc J, et al. Comparative Effectiveness of Influenza Vaccines Among US Medicare Beneficiaries Ages 65 Years and Older During the 2019-2020 Season. Clin Infect Dis. 2021;73(11):e4251–e9.

35. Faksova K, Thiesson EM, Pihlstrom N, Baum U, Biering-Sorensen T, Poukka E, et al. Brand-specific influenza vaccine effectiveness in three Nordic countries during the 2024-2025 season: a target trial emulation study based on registry data. Lancet Reg Health Eur. 2026;60:101518.

36. Zhu S, Quint J, Leon TM, Li NJ, Muldrew S, Porse C, et al. Interim Estimates of 2025-26 Seasonal Influenza Vaccine Effectiveness - California, October 2025-January 2026. MMWR Morb Mortal Wkly Rep. 2026;75(9):124–8.

37. Zhu S, Quint J, Leon TM, Sun M, Li NJ, Yen C, et al. Influenza Vaccine and Associated Infection and Death in California, 2024 to 2025. JAMA Netw Open. 2026;9(6):e2617684.

38. Zhu S, Quint J, Leon TM, Sun M, Li NJ, Yen C, et al. Estimating Influenza Vaccine Effectiveness Against Laboratory-Confirmed Influenza Using Linked Public Health Information Systems, California, 2023-2024 Season. J Infect Dis. 2025;232(5):1249–57.

39. U.S. Food & Drug Administration. Influenza Vaccine Composition for the 2025-2026 U.S. Influenza Season [Available from: https://www.fda.gov/vaccines-blood-biologics/influenza-vaccine-composition-2025-2026-us-influenza-season.

40. Centers for Disease Control and Prevention. 2025–2026 Flu Season [Available from: https://www.cdc.gov/flu/season/2025-2026.html#:∼:text=Subclade%20K%20influenza%20viruses%20have,2025%2D2026%20seasonal%20flu%20vaccines.

41. de Rooij AJH, Lempers VJC, Park Y, Vicic N, Han AX, Russell CA, et al. Reproducible and later vaccine strain selection can improve vaccine match to A/H3N2 seasonal influenza viruses. NPJ Vaccines. 2025;10(1):243.

42. Tokars JI, Olsen SJ, Reed C. Seasonal Incidence of Symptomatic Influenza in the United States. Clin Infect Dis. 2018;66(10):1511–8.

43. Somes MP, Turner RM, Dwyer LJ, Newall AT. Estimating the annual attack rate of seasonal influenza among unvaccinated individuals: A systematic review and meta-analysis. Vaccine. 2018;36(23):3199–207.

44. O’Halloran AC, Millman AJ, Holstein R, Olsen SJ, Cummings CN, Chai SJ, et al. The Burden of All-Cause Mortality Following Influenza-Associated Hospitalizations: Influenza Hospitalization Surveillance Network, 2010-2019. Clin Infect Dis. 2025;80(3):e43–e5.

45. Lomholt FK, Soborg B, Valentiner-Branth P, Slotved HC, Fuursted K, Benfield T, et al. Short-term and long-term risk of death and readmission among adults aged 50+ years admitted with severe acute respiratory infections due to COVID-19, influenza or RSV in Denmark, 2022-2025. BMJ Public Health. 2026;4(1):e004469.

