## Supplement for "The Potential Public Health Benefit of mRNA-1010 for Influenza Among Older Adults ≥65 Years in the United States"

**Supplementary Materials**

### 1 Incidence

Table 1. Overview of the CDC burden of influenza illness data used to calculate the annual incidence of influenza without vaccination

| **Flu Season** | **Population** | **Prevented Symptomatic Illnesses** | **Symptomatic Illnesses** | **Total Infections Without Vaccination** | **Seasonal Incidence** |
| --- | --- | --- | --- | --- | --- |
| 2011-2012 | 41,394,141 | 356,528 | 968,289 | 1,324,817 | 3.2% |
| 2012-2013 | 43,145,356 | 759,338 | 4,287,276 | 5,046,614 | 11.7% |
| 2013-2014 | 44,704,074 | 729,767 | 1,727,048 | 2,456,815 | 5.5% |
| 2014-2015 | 46,243,211 | 1,075,712 | 4,672,995 | 5,748,707 | 12.4% |
| 2015-2016 | 47,760,852 | 469,742 | 1,408,825 | 1,878,567 | 3.9% |
| 2016-2017 | 49,244,195 | 409,013 | 3,712,827 | 4,121,840 | 8.4% |
| 2017-2018 | 50,858,679 | 515,977 | 5,124,978 | 5,640,955 | 11.1% |
| 2018-2019 | 52,431,193 | 195,130 | 2,420,025 | 2,615,155 | 5.0% |
| 2019-2020 | 54,058,263 | 622,503 | 1,897,633 | 2,520,136 | 4.7% |
| **Total** | **429,839,964** |  |  | **31,353,606** | **7.3%** |

Figure 1. Monthly incidence of symptomatic influenza infections in the unvaccinated population, by age group (base case, plus high and low scenarios)

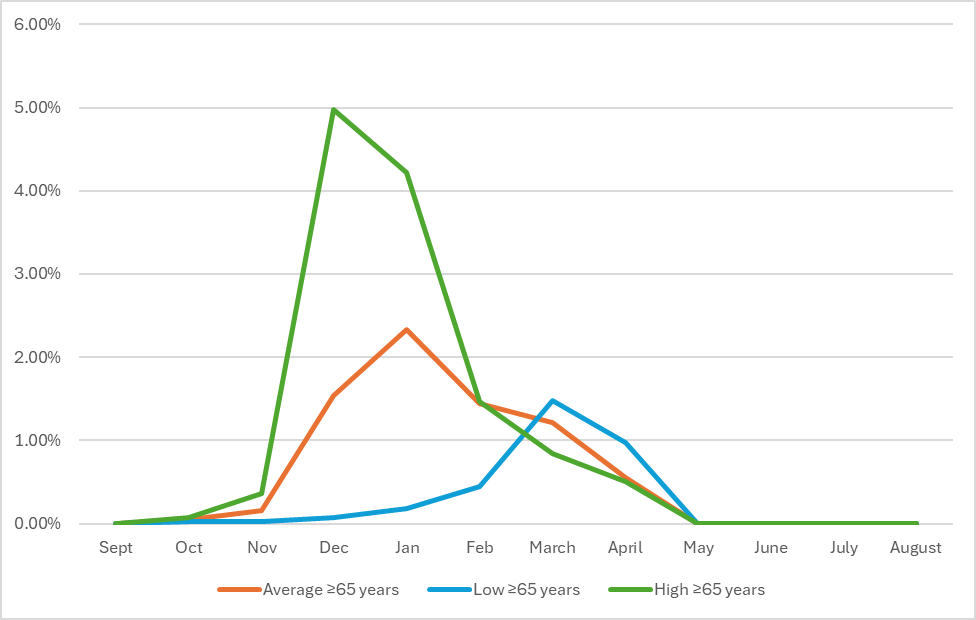

### 2 Additional Information on Vaccination

#### 2.1 Calculation of Vaccine Effectiveness

The following function was used to calculate vaccine effectiveness for each vaccine in each month of the time horizon.

$Effectiveness={Cov}_{Cur}\times{Eff}_{Cur}+{Cov}_{Cur-1}\times{Eff}_{Cur-1}+ {Cov}_{Cur-2}\times{Eff}_{Cur-2}+\ldots+ {Cov}_{Cur-9}\times{Eff}_{Cur-9}$

Where:

Cov_cur_ = fraction of age group vaccinated in the current month

Cov_cur1_ = fraction of age group vaccinated in the month before the current month

Cov_cur2_ = fraction of the age group vaccinated two months before the current month

Eff_cur_ = effectiveness in the current month

Eff_cur1_ = effectiveness one month after initial vaccination

Eff_cur2_ = effectiveness two months after initial vaccination

#### 2.2 Additional Information on the Comparator Vaccine

The seasonal inputs for VE^1^ were used to estimate initial VE (or VE in month of receipt of the vaccine) by assuming that the seasonal data represented an average VE from October to April (7 months) and that VE wanes by 8% per month.^2,3^ The solver program for optimization problems (GRG non-linear algorithm) in Microsoft Excel was used to calculate the initial VE under these constraints.

Table 2. Overview of seasonal VE and estimated initial VE used in the base case analysis^1^

| **Year** | **Seasonal VE** | **Estimated Initial VE** |
| --- | --- | --- |
| 2019-2020 | 39.0% | 63.0% |
| 2018-2019 | 12.0% | 32.8% |
| 2017-2018 | 17.0% | 39.8% |
| 2016-2017 | 40.0% | 64.0% |
| 2015-2016 | 42.0% | 66.0% |
| 2014- 2015 | 32.0% | 56.0% |
| 2013-2014 | 50.0% | 74.0% |
| 2012-2013 | 26.0% | 50.0% |
| 2011-2012 | 43.0% | 67.0% |
| *Mean* | *33.4%* | *57.0%* |
| *Standard Error* | *9.5%* | *10.3%* |

VE: Vaccine effectiveness

#### 2.3 Vaccine Coverage

Table 3. Details of vaccine coverage by month (ages 65 years and older)^4,5^

| **Year** | **Sep** | **Oct** | **Nov** | **Dec** | **Jan** | **Feb** | **Mar** | **Apr** | **May** | **Jun** | **Jul** | **Aug** |
| --- | --- | --- | --- | --- | --- | --- | --- | --- | --- | --- | --- | --- |
| **2011-2012** | 12.2% | 39.9% | 53.1% | 57.5% | 61.4% | 63.2% | 64.2% | 64.7% | 64.9% | 64.9% | 64.9% | 64.9% |
| **2012-2013** | 12.4% | 40.7% | 54.2% | 58.7% | 62.6% | 64.5% | 65.5% | 66.0% | 66.2% | 66.2% | 66.2% | 66.2% |
| **2013-2014** | 13.8% | 42.5% | 55.4% | 59.1% | 62.1% | 63.6% | 64.4% | 64.7% | 65.0% | 65.0% | 65.0% | 65.0% |
| **2014-2015** | 13.2% | 42.8% | 55.6% | 59.6% | 62.5% | 64.6% | 65.4% | 66.0% | 66.7% | 66.7% | 66.7% | 66.7% |
| **2015-2016** | 12.8% | 39.9% | 52.3% | 56.1% | 59.0% | 61.0% | 62.0% | 62.9% | 63.4% | 63.4% | 63.4% | 63.4% |
| **2016-2017** | 13.5% | 40.3% | 52.4% | 57.0% | 60.5% | 62.8% | 63.9% | 64.8% | 65.3% | 65.3% | 65.3% | 65.3% |
| **2017-2018** | 13.3% | 37.9% | 48.8% | 52.3% | 55.6% | 57.2% | 58.3% | 59.1% | 59.6% | 59.6% | 59.6% | 59.6% |
| **2018-2019** | 13.8% | 42.0% | 54.4% | 58.9% | 62.6% | 65.0% | 66.6% | 67.7% | 68.1% | 68.1% | 68.1% | 68.1% |
| **2019-2020** | 15.9% | 46.3% | 58.4% | 63.1% | 66.6% | 68.5% | 69.3% | 69.6% | 69.8% | 69.8% | 69.8% | 69.8% |
| **Average** | **13.4%** | **41.4%** | **53.8%** | **58.0%** | **61.4%** | **63.4%** | **64.4%** | **65.1%** | **65.4%** | **65.4%** | **65.4%** | **65.4%** |

### 3 Model Validation

The model was run with the average population size of the 9 seasons of interest (2011-2012 to 2019-2020), which is lower than the target population size from 2024-25. The table below compares the average clinical outcomes from the CDC burden of illness model for these 9 seasons to the model predicted number of cases with egg-based vaccination only and with no vaccination. The number of influenza cases predicted by the model was 3% higher with egg- based vaccination and >1% lower with no vaccination.

Table 4. Model validation: predicted outcomes compared to average CDC outcomes for 2011-2012 to 2019-2020

|  | **CDC Burden Model** | **Model Estimates** | **Difference** | **Percent Difference** |
| --- | --- | --- | --- | --- |
| **With Egg-based Vaccination** | | | | |
| Symptomatic Infections | 2,913,322 | 2,813,668 | 99,653 | 3% |
| Hospitalizations | 264,847 | 255,788 | 9,060 | 3% |
| Deaths | 28,017 | 27,058 | 958 | 3% |
| **With No Vaccination** | | | | |
| Symptomatic Infections | 3,483,734 | 3,482,781 | -953 | -0.03% |

### 5 Additional Result Tables and Figures

Table 5. Scenario Results

| **Scenario** | **Number of Clinical Cases Averted** | | | |
| --- | --- | --- | --- | --- |
|  | **Symptomatic Infections** | **Outpatient Treatment Only** | **Hospitalizations** | **Deaths** |
| **Base case** | 232,155 | 130,007 | 21,105 | 2,233 |
| **Vaccine effectiveness (VE) assumptions** | | | | |
| VE from COP model only | 146,562 | 82,074 | 13,324 | 1,409 |
| VE from egg adaptation only | 97,707 | 54,716 | 8,882 | 940 |
| VE assuming probability 3 of 9 for egg adaptation | 48,853 | 27,358 | 4,441 | 470 |
| VE assuming rVE of 6.5% (instead of VE from COP model) | 168,184 | 94,183 | 15,289 | 1,617 |
| **Single year** | | | | |
| 2017-2018 | 505,114 | 282,863 | 45,919 | 4,264 |
| **Incidence** | | | | |
| All seasons (2011/12-2019/20 & 2022/23-2024/25) | 217,432 | 121,762 | 19,767 | 1,992 |
| All seasons (2011/12-2019/20 & 2023/24-2024/25) | 225,518 | 126,290 | 20,502 | 2,056 |
| All seasons (2011/12-2018/19 & 2023/24-2024/25) | 243,993 | 136,636 | 22,181 | 2,228 |
| **Premature mortality costs -- include both market and non-market costs** | -- | -- | -- | -- |

Table 6. Deterministic Sensitivity Analysis Results: Clinical Outcomes

|  | **Symptomatic infections averted** | | **Hospitalizations averted** | | **Deaths averted** | |
| --- | --- | --- | --- | --- | --- | --- |
| **Sensitivity Analysis** | **Number** | **% Change from Base** | **Number** | **% Change from Base** | **Number** | **% Change from Base** |
| **Base case** | 232,155 | - | 21,105 | - | 2,233 | - |
| High incidence (2014/15) | 387,639 | 67.0% | 35,240 | 67.0% | 3,728 | 67.0% |
| Low incidence (2011/12) | 105,722 | -54.5% | 9,611 | -54.5% | 1,017 | -54.5% |
| rVE COP (95%CI UB) | 328,657 | 41.6% | 29,878 | 41.6% | 3,161 | 41.6% |
| rVE COP (95%CI LB) | 134,572 | -42.0% | 12,234 | -42.0% | 1,294 | -42.0% |
| rVE Egg Adaptation (UB; 22%) | 298,423 | 28.5% | 27,129 | 28.5% | 2,870 | 28.5% |
| rVE Egg Adaptation (LB; 2.7%) | 165,199 | -28.8% | 15,018 | -28.8% | 1,589 | -28.8% |
| rVE Egg Adaptation (probability 75%) | 242,855 | 4.6% | 22,078 | 4.6% | 2,335 | 4.6% |
| rVE Egg Adaptation (probability 50%) | 210,757 | -9.2% | 19,160 | -9.2% | 2,027 | -9.2% |
| Vaccine Effectiveness of HD-IIV (highest VE in 9 season) | 140,240 | -39.6% | 12,749 | -39.6% | 1,349 | -39.6% |
| Vaccine Effectiveness of HD-IIV (lowest VE in 9 season) | 304,367 | 31.1% | 27,670 | 31.1% | 2,927 | 31.1% |
| Coverage (Absolute +5%) - all in September | 251,013 | 8.1% | 22,819 | 8.1% | 2,414 | 8.1% |
| Coverage (Absolute -5%) - all in September | 213,298 | -8.1% | 19,391 | -8.1% | 2,051 | -8.1% |
| Probability of hospitalization (+25%) | 232,135 | 0.0% | 26,379 | 25.0% | 2,790 | 25.0% |
| Probability of hospitalization (-25%) | 232,176 | 0.0% | 15,830 | -25.0% | 1,675 | -25.0% |
| Probability of hospitalization readmissions (+25%) | 232,155 | 0.0% | 21,105 | 0.0% | 2,233 | 0.0% |
| Probability of hospitalization readmissions (-25%) | 232,155 | 0.0% | 21,105 | 0.0% | 2,233 | 0.0% |
| Mortality (High of 9 seasons) | 232,099 | 0.0% | 21,100 | 0.0% | 3,782 | 69.4% |
| Mortality (Low of 9 seasons) | 232,167 | 0.0% | 21,106 | 0.0% | 1,911 | -14.4% |

CI: Confidence Interval; COP: Correlate of protection; LB: Lower Bound; rVE: relative vaccine effectiveness; UB: Upper Bound; VE: Vaccine effectiveness

### 7 References

1.	Centers for Disease Control and Prevention. Flu burden prevented by vaccination. Accessed March 19, 2026. <https://www.cdc.gov/flu-burden/php/data-vis-vac/index.html>

2.	Ferdinands JM, Gaglani M, Martin ET, et al. Waning Vaccine Effectiveness Against Influenza-Associated Hospitalizations Among Adults, 2015-2016 to 2018-2019, United States Hospitalized Adult Influenza Vaccine Effectiveness Network. *Clin Infect Dis*. Aug 16 2021;73(4):726–729. doi:10.1093/cid/ciab045

3.	Food and Drug Administration. Influenza vaccine composition for the 2025-2026 U.S. influenza season. Accessed March 30, 2026. <https://www.fda.gov/vaccines-blood-biologics/influenza-vaccine-composition-2025-2026-us-influenza-season>

4.	Centers for Disease Control and Prevention. Influenza vaccination coverage for all ages (6+months). Accessed August 30, 2025. <https://data.cdc.gov/Flu-Vaccinations/Influenza-Vaccination-Coverage-for-All-Ages-6-Mont/vh55-3he6/about_data>

5.	Centers for Disease Control and Prevention. Weekly influenza vaccination coverage and intent for vaccination, overall, by selected demographics and jurisdictions, among adults 18 years and older. Accessed August 30, 2025. <https://data.cdc.gov/Flu-Vaccinations/Weekly-Influenza-Vaccination-Coverage-and-Intent-f/sw5n-wg2p/about_data>
